# Altered brain glucose metabolism in nicotine use but not in hazardous alcohol consumption or problem gambling of healthy middle-aged adults

**DOI:** 10.1101/2024.04.29.24306507

**Authors:** Seunghyeon Shin, Keunyoung Kim, Jihyun Kim, Hyun-Yeol Nam, Ju Won Seok, Kyoungjune Pak

## Abstract

**Objectives:** We aimed to determine whether chronic nicotine use, alcohol consumption, and gambling alters brain glucose metabolism.

**Methods:** We retrospectively analyzed data from 473 healthy men who participated in health checkups at Samsung Changwon Hospital Health Promotion Center during 2013 (baseline) and 2018 (follow-up). The health checks included a brain ^18^F-fluorodeoxyglucose positron emission tomography (PET), a questionnaire of tobacco use, the Alcohol Use Disorders Identification Test (AUDIT; Korean version), and the Problem Gambling Severity Index (PGSI). From brain PET scans, the mean uptake in regions-of-interest was scaled to the mean global cortical uptake by each individual, defining the standardized uptake value ratio. We established a model for tobacco use, AUDIT, and PGSI with regional SUVR as a dependent variable and tobacco use, AUDIT, and PGSI as predictors adjusted for age using Bayesian hierarchical modelling. Bayesian models were estimated using four Markov chains, each of which had 4,000 iterations including 1,000 warm-ups, thus totaling 12,000 post-warmup samples. The sampling parameters were slightly modified to facilitate convergence (max tree depth = 20). All data were analyzed using R (The R Foundation for Statistical Computing, Vienna, Austria).

**Results:** This study included 131 healthy males (mean age at baseline and follow-up: 43.0 ± 3.4, 48.1 ± 3.3 years, respectively). Tobacco use was negatively associated with glucose metabolism in the caudate, thalamus, cingulate, and frontal lobe, and positively associated with the cerebellum, whereas AUDIT or PGSI were not associated.

**Conclusion:** Tobacco use was associated with altered brain glucose metabolism in the caudate, thalamus, cingulate, frontal lobe, and the cerebellum. However, neither hazardous alcohol consumption, nor problem gambling showed any association with brain glucose metabolism. Our findings might provide new insights into the neural mechanisms of chronic nicotine use.

## INTRODUCTION

The global number of smokers worldwide during 2019 was 1.14 billion^1^. The age-standardized prevalence of current tobacco smokers was 32.7% among men aged over 15 years and 6.62% among women^1^. Although the prevalence of smoking had decreased significantly since 1990, the total number of smokers has increased^1^. Tobacco smoking is responsible for the premature deaths of approximately 6 million people annually worldwide ^2^. Most smoking-related deaths arise from cancers, respiratory, and cardiovascular diseases^2^. Smoking is a risk factor for stroke, blindness, deafness, back pain, osteoporosis, and peripheral vascular disease^2^.

Cigarette addiction is derived from nicotine in tobacco^2^ that enters the bloodstream through the lungs and reaches the brain across the blood-brain barrier^3^. This results in elevated blood pressure, glucose release, increased respiration, heart rates, and alertness, as well as constricted arteries^3^.

Acetylcholine plays a role in mental and physical arousal, learning, memory, and emotion and binds to n-acetylcholine receptors (nAChRs) that are classified as nicotine and muscarine types^3^. Nicotine also binds to nAChRs that are localized throughout the brain and peripheral nervous system including the midbrain, striatum, nucleus accumbens (NAc), ventral tegmentum, muscle, adrenal glands, and heart^4^. The gratifying action of nicotine is caused by the activation of dopaminergic neurons in the ventral tegmental area that projects to the NAc and prefrontal cortex^4^. Nicotine administration increases dopaminergic neuron firing in striatal and extrastriatal areas resulting in elevated dopamine levels^4^. The effects of smoking on the brain have been investigated in detail. The volumes of grey and white matter are decreased in magnetic resonance imaging (MRI) scans of smokers compared with non-smokers^5-7^. According to a meta-analysis, activation of the striatum and anterior cingulate gyrus increases in response to smoking-related *vs*. neutral cues in smokers^8^. Also, connectivity is lower in the orbitofrontal cortex, superior frontal gyrus, temporal lobe and insula in smokers compared with non-smokers^9^. However, the chronic effects of nicotine on brain glucose metabolism have not been evaluated. Thus, we aimed to identify the effects of chronic nicotine inhalation measured as pack-years on brain glucose metabolism. We also evaluated correlations between brain glucose metabolism and alcohol consumption as well as gambling, since these activities decrease grey matter volume^10,11^.

## METHODS

### Participants

We retrospectively analyzed data from 473 healthy men who participated in health check-ups at Samsung Changwon Hospital Health Promotion Center during 2013 (baseline) and 2018 (follow-up). We excluded individuals with neuropsychiatric disorders (n = 5), malignancies (n = 3), as well as missing tobacco use, Alcohol Use Disorders Identification Test (AUDIT; Korean version), and Problem Gambling Severity Index (PGSI) data (n = 302). We finally analyzed data from 131 healthy men. The health checks included a brain ^18^F-FDG positron emission tomography (PET), questionnaires about tobacco consumption, AUDIT, and PGSI. All 131 men had been included in a previous study of the effects of aging on brain glucose metabolism^12^. The Institutional Review Board of Changwon Samsung Hospital approved the study protocol and the need for informed consent to participate was waived due to the retrospective study design.

### Brain ^18^F-FDG PET and Image analysis

The participants avoided strenuous exercise for 24 hours and fasted for at least 6 hours before starting the PET study. The PET/computed tomography (CT) images were acquired from 60 minutes after injecting the participants with^18^F-FDG (3.7 MBq/kg) using a Discovery 710 PET/CT scanner (GE Healthcare, Waukesha, WI, USA). Continuous spiral CT images were acquired under the parameters of tube voltage and current of 120 kVp and 30-180 mAs, respectively. The PET images were acquired in 3-dimensional mode with an FWHM of 5.6 mm and reconstructed using an ordered-subset expectation maximization (OSEM) algorithm. The images were spatially normalized to Montreal Neurological Institute (MNI) space using PET templates from Statistical Parametric Mapping 5 (SPM 5; (University College of London, UK) with the reference tool for PET kinetic modeling pmod v. 3.6 (PMOD Technologies LLC, Zurich, Switzerland). Regions of interest (ROIs) was defined using the Automated Anatomical Labeling 2 (AAL2) atlas^13^, which comprised the caudate, cerebellum, cingulate, frontal lobe, hippocampus, insula, occipital lobe, pallidum, parietal lobe, putamen, temporal lobe, and thalamus. The mean uptake of each ROI was scaled to the mean global cortical uptake of each individual, and defined as the standardized uptake value ratio (SUVR). We smoothed SUVR images with a Gaussian kernel of FWHM 8 mm using Statistical Parametric Mapping (SPM) 12 (Wellcome Centre for Human Neuroimaging, UCL, London, UK). The statistical threshold was set at the cluster level for full-volume analysis and corrected using a false discovery rate with p < 0.05 in a regression model (corrected for age).

### Tobacco use, AUDIT, and PGSI

Tobacco use was analyzed in terms of pack-years by multiplying the number of packs of cigarettes smoked daily by the number of years. The participants were screened using AUDIT for hazardous consumption or any alcohol use disorder. It consists of 10 questions that are each scored from 0 to 4 with higher scores indicating increased risk of alcohol use disorder. Scores from 0□7, 8□14, and ≥ 15 respectively indicate low-risk, hazardous or harmful alcohol consumption, and a likelihood of alcohol dependence^14^. PGSI is a Korean version of Canadian Problem Gambling Index^15^ that screens for gambling problems. It consists of nine questions that are each scored from 0□3, with higher scores indicating increased risk of problem gambling. Scores of 0, 1□2, 3□7, and ≥8 indicate a gambler without a problem, and low-risk, moderate-risk and problem gamblers, respectively.

### Statistical analysis

Normality was assessed using Shapiro–Wilk tests. After logarithmic transformation of the regional SUVR, we analyzed the effects of chronic tobacco use, alcohol consumption (AUDIT), and gambling (PGSI) on regional SUVRs using Bayesian hierarchical modelling with brms^16-18^ that applies Markov-Chain Monte Carlo sampling using the rstan package in R^19^. We established a model for tobacco use, AUDIT, and PGSI with regional SUVR as a dependent variable and tobacco use, AUDIT, and PGSI as predictors adjusted for age. These fixed effects were individually calculated and participants and ROIs were added as random intercepts to allow the SUVR to vary between them. Bayesian models were estimated using four Markov chains, each of which had 4,000 iterations including 1,000 warm-ups, thus totaling 12,000 post-warmup samples. The sampling parameters were slightly modified to facilitate convergence (max tree depth = 20). All data were analyzed using R (The R Foundation for Statistical Computing, Vienna, Austria).

## RESULTS

### Characteristics of participants

This study included 131 healthy men (mean age at baseline and follow-up: 43.0 ± 3.4, 48.1 ± 3.3 years, respectively). The mean and standard deviations of tobacco use (pack-years), AUDIT, and PGSI were 15.5 ± 11.4, 11.4 ± 6.1, 0.3 ± 0.9 at baseline, 15.1 ± 10.6, 10.4 ± 6.1, 0.2 ± 1.1 at follow-up.

### Tobacco use, AUDIT, PGSI, and Brain glucose metabolism

Tobacco use was negatively associated with glucose metabolism in the caudate, thalamus, cingulate, and frontal lobe, and positively with that in the cerebellum at baseline and follow-up (Figure 1). Full-volume analysis consistently revealed a negative association between tobacco use and glucose metabolism in the caudate, thalamus, cingulate, and frontal lobe, and a positive association with the cerebellum (Figure 2). However, Bayesian models and full-volume analyses did not associate brain glucose metabolism with either AUDIT or PGSI.

**Figure 1.**
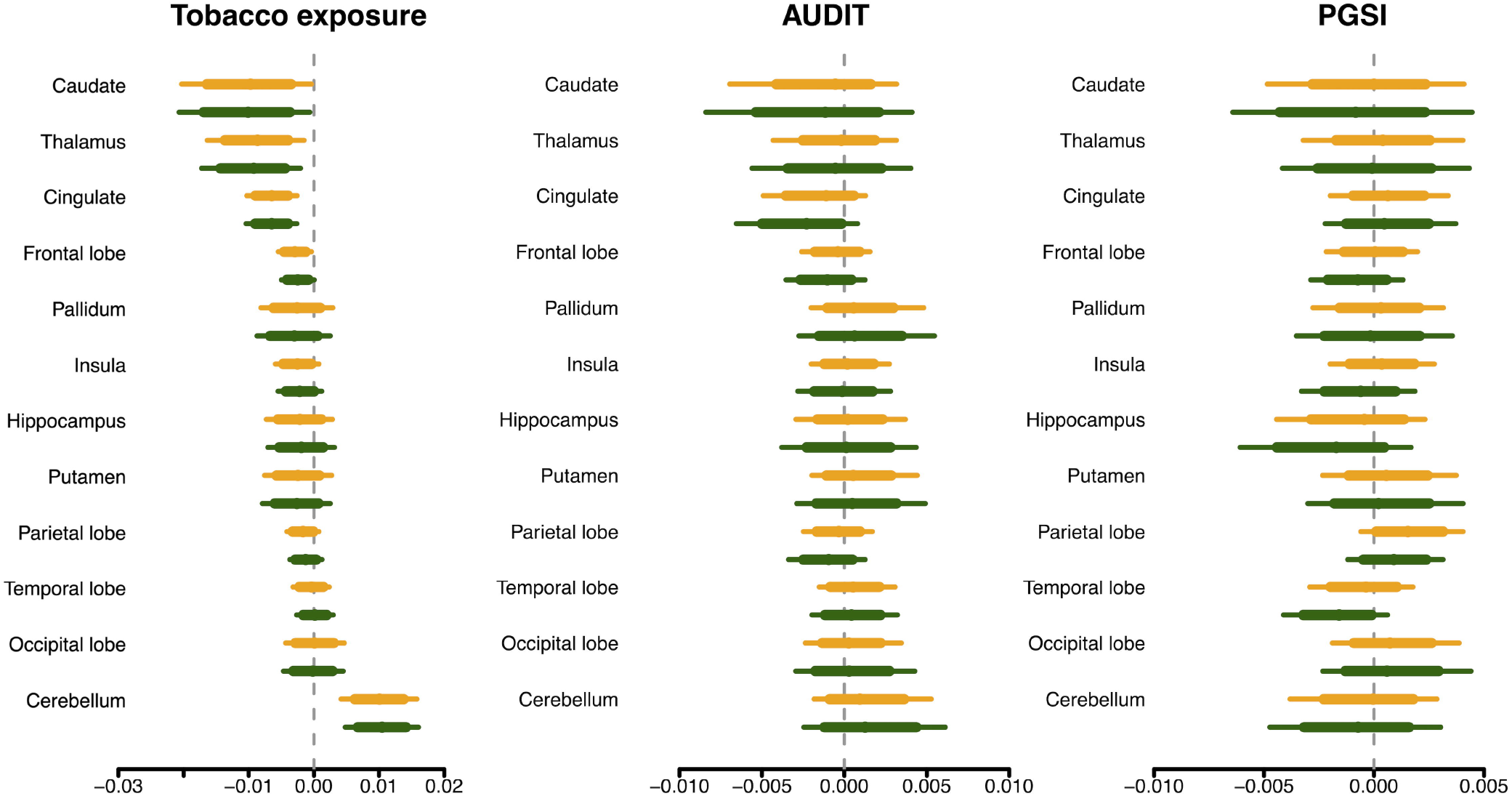
Posterior intervals of regression coefficients for tobacco use, AUDIT and PGSI scores predicting brain glucose metabolism. Thick and thin lines represent 80% and 95% posterior intervals, respectively. Orange, baseline; green, follow-up. AUDIT, Alcohol Use Disorders Identification Test; PGSI, Problem Gambling Severity Index.

**Figure 2.**
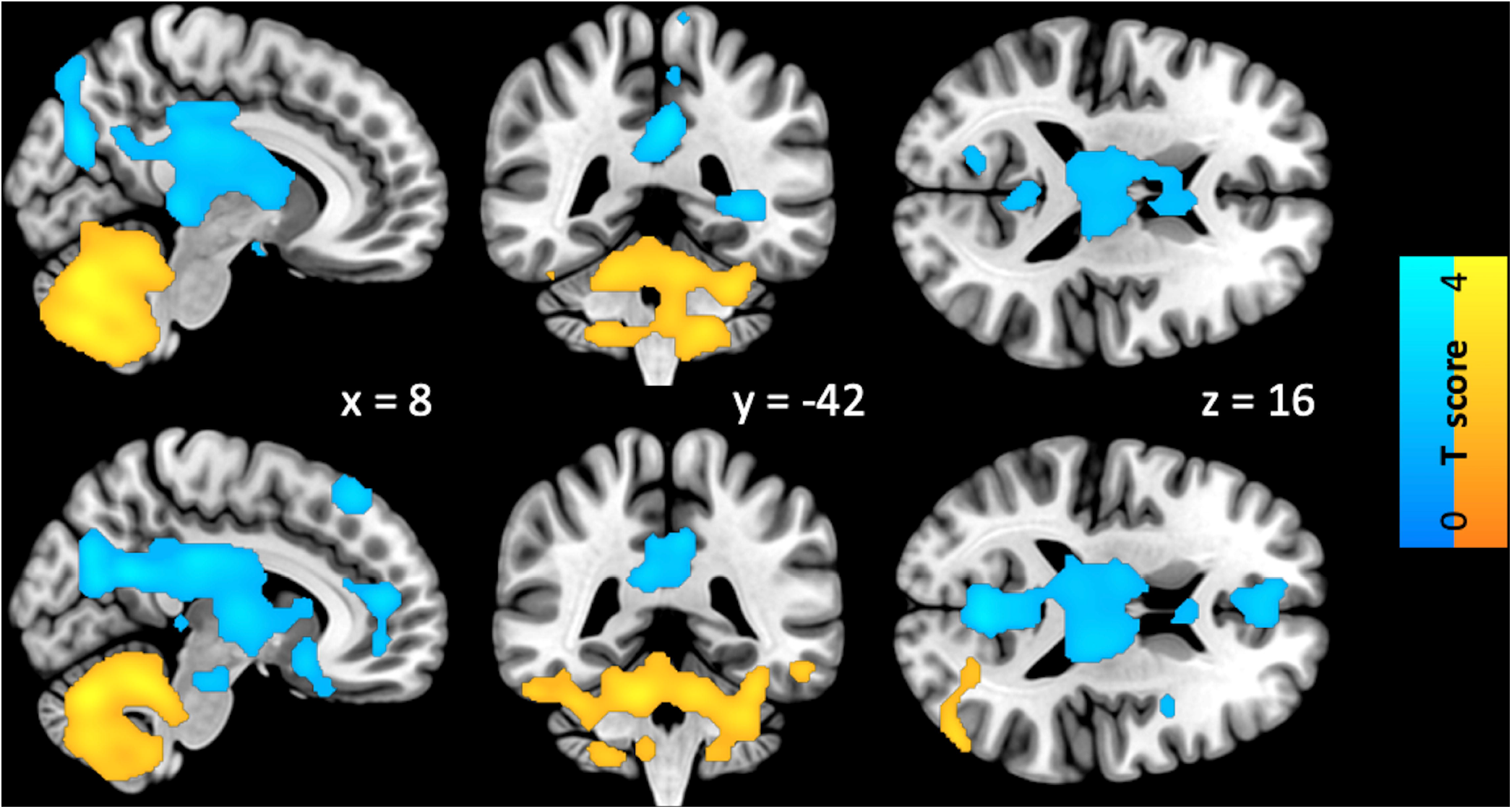
Full volume analysis of association between tobacco use and brain glucose metabolism. Upper and lower rows, baseline and follow-up studies, respectively. Yellow and blue, positive and negative associations, respectively.

## DISCUSSION

Our main finding was that tobacco use was associated with altered brain glucose metabolism in the caudate, thalamus, cingulate, frontal lobe, and the cerebellum. However, neither hazardous alcohol consumption, nor problem gambling showed any association with brain glucose metabolism. These effects are also consistent over the studied period of 5 years in the middle adulthood.

The caudate nucleus plays a role in the establishment and expression of reward-related conditioned behavior^20^, and caudate reactivity to smoking cues is associated with a reward response ^20^. An increased caudate volume in young male smokers has been identified using MRI^21^, whereas others found that young male smokers had a smaller caudate volume^22^. The authors speculated that the difference might be due to nicotine dependence and the consequent accumulation of neurotoxic effects^22^. The thalamus expresses abundant nAChR^23^. Acute administration of nicotine activates the thalamus, and chronic nicotine use leads to decreased nAChR in the thalamus ^24^. The thalamus plays a role in drug addiction *via* a locomotor response to psychostimulants, cue-reward association, the expression of drug-induced place preference, withdrawal, and the reinstatement of drug-seeking^25^. Individuals who are dependent on alcohol, cocaine, nicotine, methamphetamine, opioids, cannabis, and synthetic cannabinoids have lower thalamic volumes^26^. Thalamic connectivity is also decreased in those who are dependent on alcohol, cocaine, and nicotine^26^. A meta-analysis of chronic cigarette smoking found a decreased volume of grey matter in the thalamus^27^. The cingulate cortex expresses nAChR and is associated with nicotine^28,29^. The cingulate gyrus of smokers becomes activated in response to smoking, rather than neutral cues^30^. The cingulate cortex volume is smaller among smokers compared with nonsmokers^5,7,31,32^. A decreased cingulate cortex volume is also associated with alcohol, methamphetamine, and cocaine dependance^33^. An increased severity of nicotine addiction is associated with weaker functional connectivity between the cingulate cortex and striatum^28^. Nicotine also affects the frontal cortex. Grey matter volume and frontal cortex density are decreased in smokers^5,32^. The volume of the frontal cortex negatively correlates with pack-years ^6^. Nicotine is associated with disturbed working memory and attention that correlate with the prefrontal cortex, the activity of which is reduced^34,35^. The cerebellum also functions in addiction^36^. The cerebellum correlates with functional resting state networks including the prefrontal cortex, cingulate, and insula that are associated with cognitive control, stimulus salience, and interoception, and shares reciprocal connections with the dopaminergic system in basal ganglia^36^. Addictive cocaine, opioid, nicotine, and alcohol are associated with decreased grey matter volume^36^. Smokers have smaller cerebellar grey matter volumes, reduced grey matter density^5,7,37,38^, and functional connectivity^38^. The neurotoxic effect of nicotine use might be responsible for the decreased volume, density, and connectivity in grey matter ^27,37^. These effects might appear as negative correlations between pack-years and brain glucose metabolism measured by ^18^F-FDG PET in the caudate, thalamus, cingulate, and frontal lobe. However, the correlation between pack-years and brain glucose metabolism in cerebellum was positive in the present study. The cerebellum has nAChRs that are sensitive to nicotine, which increases both glutamatergic GABAergic transmission by binding to nAChRs, but the former is superior to the latter^39^. Chronic nicotine use increases glucose oxidation and neurotransmitter cycling by glutamatergic neurons in the cortex, subcortex and cerebellum^40^. However, GABAergic metabolism increases in the cortex and but not in the subcortical and cerebellum regions^40^. Thus, chronic nicotine use increases excitatory glutamatergic transmission in the cerebellum^39^. Furthermore, cerebellar glucose metabolism positively correlates with age in healthy persons^41,42^. Age-related degeneration of the cerebellum is assumed to start between the ages of 50 and 60 years, and its age-related volume loss is lower than that of the cerebrum^43^. The aging effect on the cerebellum and increased glucose oxidation due to chronic nicotine use might explain the positive correlation regardless of a decreased cerebellar volume. Further investigation is needed to validate this.

The effects of alcohol on the brain are established. Alcohol consumption negatively correlates with total cerebral brain and grey matter volumes^10,44^, even in middle aged individuals^45^. The findings of ^18^F-FDG PET imaging associated alcohol use disorder with decreased glucose metabolism in the frontal, temporal, and parietal lobes compared with age- and sex-matched healthy persons^46^. The present study did not identify a correlation between AUDIT scores and brain glucose metabolism. However, AUDIT scores do not include the duration of alcohol consumption, and high scores do not necessarily indicate alcohol use disorder. Thus, despite a higher AUDIT score, changes in the brain might not be eminent. Behavioral addictions, including pathological gambling, decrease grey matter volumes compared with healthy controls^11^. Relative glucose metabolic rates in the orbitofrontal and medial frontal cortex are significantly increased compared with healthy controls^47^.

The present study did not identify a correlation between the PGSI score and brain glucose metabolism.

The PGSI score of most participants in this study was 0, which might be responsible for the absence of a correlation between PGSI scores and brain glucose metabolism.

This retrospective study has several limitations. Because we included only men, our results might not be directly generalized to women. The study was based on a health check program that did not include brain MRI. Thus, MRI-based coregistration and partial volume correction of PET images were impossible and the results could not be compared with MRI-based indices of atrophy.

## CONCLUSION

Tobacco use was associated with altered brain glucose metabolism in the caudate, thalamus, cingulate, frontal lobe, and the cerebellum. However, neither hazardous alcohol consumption, nor problem gambling showed any association with brain glucose metabolism. Our findings might provide new insights into the neural mechanisms of chronic nicotine use.

## Data Availability

All data produced in the present study are available upon reasonable request to the authors

## TABLES

**Table.** Subjects’ characteristics.

|  | Baseline | Follow-up | p |
| --- | --- | --- | --- |
| Age (years) | 43.0±3.4 | 48.1±3.3 | <0.0001 |
| Body mass index (kg/m<sup>2</sup>) | 24.8±2.8 | 24.7±2.7 | 0.6553 |
| Tobacco exposure (pack year) | 15.5±11.4 | 15.1±10.6 | 0.6375 |
| Alcohol Use Disorders Identification Test | 11.4±6.1 | 10.4±6.1 | 0.0054 |
| 0~7 (low risk) | 34 | 45 |  |
| 8~14 (hazardous or harmful range) | 60 | 58 |  |
| 15~ (alcohol dependent range) | 37 | 28 |  |
| Problem Gambling Severity Index | 0.3±0.9 | 0.2±1.1 | 0.5387 |
| 0 (non-problem gambler) | 115 | 124 |  |
| 1~2 (low-risk gambler) | 11 | 4 |  |
| 3~7 (moderate-risk gambler) | 5 | 1 |  |
| 8~ (problem gambler) | 0 | 2 |  |

## REFERENCES

1. Collaborators GBDT. Spatial, temporal, and demographic patterns in prevalence of smoking tobacco use and attributable disease burden in 204 countries and territories, 1990-2019: a systematic analysis from the Global Burden of Disease Study 2019. Lancet. 2021;397(10292):2337–2360.

2. West R. Tobacco smoking: Health impact, prevalence, correlates and interventions. Psychol Health. 2017;32(8):1018–1036.

3. Jiloha RC. Biological basis of tobacco addiction: Implications for smoking-cessation treatment. Indian J Psychiatry. 2010;52(4):301–307.

4. Le Foll B, Piper ME, Fowler CD, et al. Tobacco and nicotine use. Nat Rev Dis Primers. 2022;8(1):19.

5. Brody AL, Mandelkern MA, Jarvik ME, et al. Differences between smokers and nonsmokers in regional gray matter volumes and densities. Biol Psychiatry. 2004;55(1):77–84.

6. Peng P, Li M, Liu H, et al. Brain Structure Alterations in Respect to Tobacco Consumption and Nicotine Dependence: A Comparative Voxel-Based Morphometry Study. Front Neuroanat. 2018;12:43.

7. Peng P, Wang Z, Jiang T, Chu S, Wang S, Xiao D. Brain-volume changes in young and middle-aged smokers: a DARTEL-based voxel-based morphometry study. Clin Respir J. 2017;11(5):621–631.

8. Lin X, Deng J, Shi L, et al. Neural substrates of smoking and reward cue reactivity in smokers: a meta-analysis of fMRI studies. Transl Psychiatry. 2020;10(1):97.

9. Zhou S, Xiao D, Peng P, et al. Effect of smoking on resting-state functional connectivity in smokers: An fMRI study. Respirology. 2017;22(6):1118–1124.

10. Topiwala A, Ebmeier KP, Maullin-Sapey T, Nichols TE. Alcohol consumption and MRI markers of brain structure and function: Cohort study of 25,378 UK Biobank participants. Neuroimage Clin. 2022;35:103066.

11. Qin K, Zhang F, Chen T, et al. Shared gray matter alterations in individuals with diverse behavioral addictions: A voxel-wise meta-analysis. J Behav Addict. 2020;9(1):44–57.

12. Pak K, Malen T, Santavirta S, et al. Brain Glucose Metabolism and Aging: A 5-Year Longitudinal Study in a Large Positron Emission Tomography Cohort. Diabetes Care. 2023;46(2):e64–e66.

13. Rolls ET, Joliot M, Tzourio-Mazoyer N. Implementation of a new parcellation of the orbitofrontal cortex in the automated anatomical labeling atlas. Neuroimage. 2015;122:1–5.

14. Saunders JB, Aasland OG, Babor TF, de la Fuente JR, Grant M. Development of the Alcohol Use Disorders Identification Test (AUDIT): WHO Collaborative Project on Early Detection of Persons with Harmful Alcohol Consumption--II. Addiction. 1993;88(6):791–804.

15. Ah-Young Kim, Jungeun Cha, Sun Jung Kwon, Soonmook Lee. Construction and Validation of Korean Version of CPGI. Korean Journal of Psychology: General. 2011;30(4):1011–1038.

16. Bürkner P-C. Bayesian Item Response Modeling in R with brms and Stan. Journal of Statistical Software. 2021;100(5):1–54.

17. Bürkner P-C. brms: an R package for Bayesian multilevel models using Stan. Journal of Statistical Software. 2017;80(1):1–28.

18. Bürkner P-C. Advanced Bayesian multilevel modeling with the R package brms. The R Journal. 2018;10(1):395–411.

19. Stan Development Team. RStan: the R interface to Stan. https://mc-stan.org/. Published 2022. Accessed.

20. Molokotos E, Peechatka AL, Wang KS, Pizzagalli DA, Janes AC. Caudate reactivity to smoking cues is associated with increased responding to monetary reward in nicotine-dependent individuals. Drug Alcohol Depend. 2020;209:107951.

21. Li Y, Yuan K, Cai C, et al. Reduced frontal cortical thickness and increased caudate volume within fronto-striatal circuits in young adult smokers. Drug Alcohol Depend. 2015;151:211–219.

22. Lin F, Han X, Wang Y, et al. Sex-specific effects of cigarette smoking on caudate and amygdala volume and resting-state functional connectivity. Brain Imaging Behav. 2021;15(1):1–13.

23. Majdi A, Kamari F, Vafaee MS, Sadigh-Eteghad S. Revisiting nicotine’s role in the ageing brain and cognitive impairment. Rev Neurosci. 2017;28(7):767–781.

24. Swan GE, Lessov-Schlaggar CN. The effects of tobacco smoke and nicotine on cognition and the brain. Neuropsychol Rev. 2007;17(3):259–273.

25. De Groote A, de Kerchove d’Exaerde A. Thalamo-Nucleus Accumbens Projections in Motivated Behaviors and Addiction. Front Syst Neurosci. 2021;15:711350.

26. Huang AS, Mitchell JA, Haber SN, Alia-Klein N, Goldstein RZ. The thalamus in drug addiction: from rodents to humans. Philos Trans R Soc Lond B Biol Sci. 2018;373(1742).

27. Sutherland MT, Riedel MC, Flannery JS, et al. Chronic cigarette smoking is linked with structural alterations in brain regions showing acute nicotinic drug-induced functional modulations. Behav Brain Funct. 2016;12(1):16.

28. Hong LE, Gu H, Yang Y, et al. Association of nicotine addiction and nicotine’s actions with separate cingulate cortex functional circuits. Arch Gen Psychiatry. 2009;66(4):431–441.

29. Pichon S, Garibotto V, Wissmeyer M, et al. Higher availability of alpha4beta2 nicotinic receptors (nAChRs) in dorsal ACC is linked to more efficient interference control. Neuroimage. 2020;214:116729.

30. Engelmann JM, Versace F, Robinson JD, et al. Neural substrates of smoking cue reactivity: a meta-analysis of fMRI studies. Neuroimage. 2012;60(1):252–262.

31. van de Weijer MP, Vermeulen J, Schrantee A, Munafo MR, Verweij KJH, Treur JL. The potential role of gray matter volume differences in the association between smoking and depression: A narrative review. Neurosci Biobehav Rev. 2024;156:105497.

32. Fritz HC, Wittfeld K, Schmidt CO, et al. Current smoking and reduced gray matter volume-a voxel-based morphometry study. Neuropsychopharmacology. 2014;39(11):2594–2600.

33. Hill-Bowen LD, Riedel MC, Salo T, et al. Convergent gray matter alterations across drugs of abuse and network-level implications: A meta-analysis of structural MRI studies. Drug Alcohol Depend. 2022;240:109625.

34. Lara AH, Wallis JD. The Role of Prefrontal Cortex in Working Memory: A Mini Review. Front Syst Neurosci. 2015;9:173.

35. Goriounova NA, Mansvelder HD. Short- and long-term consequences of nicotine exposure during adolescence for prefrontal cortex neuronal network function. Cold Spring Harb Perspect Med. 2012;2(12):a012120.

36. Moulton EA, Elman I, Becerra LR, Goldstein RZ, Borsook D. The cerebellum and addiction: insights gained from neuroimaging research. Addict Biol. 2014;19(3):317–331.

37. Gallinat J, Meisenzahl E, Jacobsen LK, et al. Smoking and structural brain deficits: a volumetric MR investigation. Eur J Neurosci. 2006;24(6):1744–1750.

38. Shen Z, Huang P, Wang C, Qian W, Yang Y, Zhang M. Cerebellar Gray Matter Reductions Associate With Decreased Functional Connectivity in Nicotine-Dependent Individuals. Nicotine Tob Res. 2018;20(4):440–447.

39. Wang YB, Lan Y. The Role of the Cerebellum in Drug Reward: A Review. J Integr Neurosci. 2023;22(6):147.

40. Shameem M, Patel AB. Glutamatergic and GABAergic metabolism in mouse brain under chronic nicotine exposure: implications for addiction. PLoS One. 2012;7(7):e41824.

41. Subtirelu RC, Teichner EM, Su Y, et al. Aging and Cerebral Glucose Metabolism: (18)F-FDG-PET/CT Reveals Distinct Global and Regional Metabolic Changes in Healthy Patients. Life (Basel). 2023;13(10).

42. Allocca M, Linguanti F, Calcagni ML, et al. Evaluation of Age and Sex-Related Metabolic Changes in Healthy Subjects: An Italian Brain 18F-FDG PET Study. J Clin Med. 2021;10(21).

43. Luft AR, Skalej M, Schulz JB, et al. Patterns of age-related shrinkage in cerebellum and brainstem observed in vivo using three-dimensional MRI volumetry. Cereb Cortex. 1999;9(7):712–721.

44. Paul CA, Au R, Fredman L, et al. Association of alcohol consumption with brain volume in the Framingham study. Arch Neurol. 2008;65(10):1363–1367.

45. Immonen S, Launes J, Jarvinen I, et al. Moderate alcohol use is associated with decreased brain volume in early middle age in both sexes. Sci Rep. 2020;10(1):13998.

46. Bralet MC, Mitelman SA, Goodman CR, Lincoln S, Hazlett EA, Buchsbaum MS. Fluorodeoxyglucose positron emission tomography scans in patients with alcohol use disorder. Alcohol Clin Exp Res. 2022;46(6):994–1010.

47. Hollander E, Buchsbaum MS, Haznedar MM, et al. FDG-PET study in pathological gamblers. Lithium increases orbitofrontal, dorsolateral and cingulate metabolism. Neuropsychobiology. 2008;58(1):37–47.

